# Shear-Wave Anisotropy of the Vastus Lateralis During Low-Level Isometric Contraction Measured with Ultrasound Time-Harmonic Elastography

**DOI:** 10.1101/2025.09.30.25336813

**Authors:** Tom Meyer, Stefan Klemmer Chandía, Pascal Engl, Giacomo Valli, Yanglei Wu, Klaus Jenderka, Thomas Bartels, René Schwesigd, Jing Guo, Eduard Kurz, Ingolf Sack, Hossein S. Aghamiry

## Abstract

**Purpose:** Skeletal muscle is commonly modeled as a transverse isotropic medium, however, the behavior of its anisotropy under active loading remains insufficiently characterized. In this study we used ultrasound time-harmonic elastography (THE) to quantify direction-dependent shear-wave speed (SWS) in the vastus lateralis (VL) muscle at rest and during low isometric contraction intensities.

**Methods:** Twenty-six healthy adults (15 men, 11 women; **25.0 *±* 4.1** y) underwent multi-frequency THE (60–80 Hz). The transducer was aligned parallel (longitudinal) and perpendicular (transverse) to VL fascicles, and measurements were acquired at rest and at 15% and 30% of maximal voluntary contraction (MVC). The anisotropy index (AI) was defined as ***AI* = *SW S***_**∥**_***/SW S***_**⊥**_. Orientation and contraction effects were tested with repeated-measures analyses.

**Results:** At rest, longitudinal SWS exceeded transverse SWS (**2.5 *±* 0.2** vs. **1.4 *±* 0.1** m/s; paired t-test ***p <* 0.01**). With contraction, SWS increased to **3.2 *±* 0.2** and **3.8 *±* 0.3** m/s (15%, 30% MVC) along fibers, and to **1.6 *±* 0.1** and **1.8 *±* 0.1** m/s across fibers (all ***p <* 0.01**). A two-factor repeated-measures ANOVA on SWS showed main effects of orientation and contraction and a significant interaction (all ***p <* 0.01**). AI increased from **1.7 *±* 0.1** at rest to **2.0 *±* 0.1** at 15% and **2.1 *±* 0.1** at 30% MVC (***p <* 0.01**). No sex- or BMI-related effects were detected.

**Conclusion:** VL exhibited marked shear-wave anisotropy at rest that increased with low-level contraction intensities, indicating disproportionate stiffening along the fiber direction. THE provides a rapid, cost-effective, orientation-sensitive readout of muscle mechanics that may support studies of neuromuscular function and pathology.

## 1 Introduction

Skeletal muscle is architecturally complex and mechanically anisotropic, with material properties that differ markedly along versus across fibers [1–4]. Force is transmitted primarily along the fiber direction [5], while the extracellular matrix channels load across fibers [6]. Quantifying in vivo anisotropy, the direction-dependent shear stiffness measured along and across muscle fibers, is essential for interpreting normal function [7], adaptations to training or disuse [8, 9], and a wide range of neuromuscular and musculoskeletal conditions [10].

Shear-wave–based elastography provides a non-invasive window into muscle mechanics [11–15]. In its simplest use, tissue is treated as isotropic and quasi-incompressible, relating shear-wave speed (SWS) to the shear modulus *µ* via *µ* = *ρ* SWS^2^. This approximation is useful in organs such as liver or breast, but it fails in skeletal muscle, whose organized fibers and connective tissue impart transverse isotropy (TI) on a macroscopic level. This means shear waves travel faster along fibers than across them [1, 14, 16–18]. Ignoring anisotropy can therefore confound interpretation and apparent “stiffness changes” may instead reflect probe orientation or wave-field directionality [3, 19].

Beyond measurement accuracy, anisotropy itself is biologically meaningful. TI-aware modeling of incompressible materials separates three independent parameters—*µ*_∥_, *µ*_⊥_, and the ratio of Young’s moduli parallel and perpendicular to the fiber direction [19–21]. This separation provides direction-dependent stiffness needed for realistic force estimation and offers biomarkers that may change with architecture (aging, disuse, spasticity, training) or pathology even when scalar stiffness is only modestly altered [3, 13, 14, 22].

Magnetic resonance elastography (MRE) has demonstrated that incorporating TI improves separation of healthy and pathological muscle and enables multiparametric characterization with multi-frequency data [21, 23, 24]. However, the cost, post-processing burden, and limited availability of MRE constrain routine use. Ultrasound-based shear-wave elastography (SWE) offers a pragmatic alternative with bedside accessibility; ultrafast imaging, rotational three-dimensional shear-wave elasticity imaging (SWEI) [3], and Viscoelastic Response (VisR) methods allow real-time, direction-specific assessment in vivo [22].

Within this ultrasound family, time-harmonic elastography (THE) [15, 25, 26] is particularly cost-effective because it uses low-amplitude external vibration and standard line-by-line acquisition, avoiding high-power acoustic radiation force impulse (ARFI) pushes and ultrafast plane-wave hardware.

Here we use THE to quantify direction-dependent SWS in the vastus lateralis (VL) muscle at rest and during low-level isometric contractions. Because externally driven fields contain mixtures of propagation directions and polarizations, we applied narrowband directional filtering and identical processing to acquisitions with the transducer aligned parallel (longitudinal) and perpendicular (transverse) to VL fibers prior to inversion. We report anisotropy based on the index *AI* = SWS_∥_*/*SWS_⊥_ (subscripts ∥ and ⊥ denote SWS measured along and perpendicular to the fascicle direction, respectively) and, given the pennate architecture of VL, estimated the in-plane fascicle angle and applied a small-angle correction to minimize orientation bias.

Our objectives were to (i) quantify VL anisotropy at rest, (ii) test how SWS and *AI* change from rest to 15% and 30% maximal voluntary contraction (MVC) intensities, and (iii) assess whether muscle contraction affects anisotropy. We hypothesized that SWS would increase with contraction level in both orientations and that anisotropy would be maintained or increase due to disproportionate stiffening along fibers.

## 2 Materials and Methods

### 2.1 Participants

Twenty-six healthy adults (15 men, 11 women) were included in this study (Table 1). All participants were free of lower-limb injury and neuromuscular disease and refrained from vigorous physical activity for 24 h before testing. The study was approved by the local ethics committee (Berlin: EA4/040/22, Halle: 2024-208) and conducted in accordance with the Declaration of Helsinki. Written informed consent was obtained from all participants prior to enrollment.

**Table 1.**
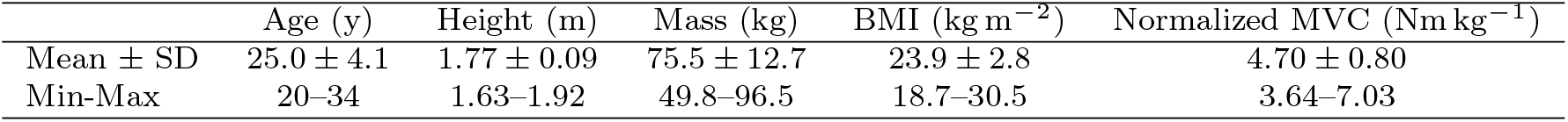
Demographic characteristics of the study cohort (*N* = 26). Here, BMI denotes Body-mass index. Also, the normalized MVC was computed by multiplying each participant’s maximum MVC by leg length to obtain knee-extension torque and then dividing by body mass, yielding a mass-normalized maximal voluntary torque reported in Nm kg^−1^.

| | Age (y) | Height (m) | Mass (kg) | BMI ($\text{kg m}^{-2}$) | Normalized MVC ($\text{Nm kg}^{-1}$) |
| --- | --- | --- | --- | --- | --- |
| Mean ± SD | $25.0 \pm 4.1$ | $1.77 \pm 0.09$ | $75.5 \pm 12.7$ | $23.9 \pm 2.8$ | $4.70 \pm 0.80$ |
| Min-Max | 20–34 | 1.63–1.92 | 49.8–96.5 | 18.7–30.5 | 3.64–7.03 |

### 2.2 Experimental setup

SWS in VL was measured with THE in two transducer orientations—longitudinal (parallel to fibers) and transverse (perpendicular to fibers)—following the anisotropy protocol of Lima *et al*. [2]. Participants were seated upright with the right leg supported, the hip at ≈90^*°*^ and the knee at 90^*°*^ (Fig. 1a). A THE device (GAMPT, Merseburg, Germany) with a linear ultrasound transducer (5–11 MHz, 54.6 mm footprint; LF11-5H60-A3, TeleMed) was positioned over VL at 75% of the distance between the greater trochanter and the lateral femoral epicondyle. The transducer was secured using an elastic strap and a custom 3D-printed holder that allowed rotation in 22.5^*°*^ increments (Fig. 1b), minimizing movement and operator variability and enabling precise 90^*°*^ rotations between longitudinal and transverse measurements. The initial longitudinal position was marked on the skin to ensure consistent placement before rotating four steps to obtain the transverse orientation.

**Fig. 1.**
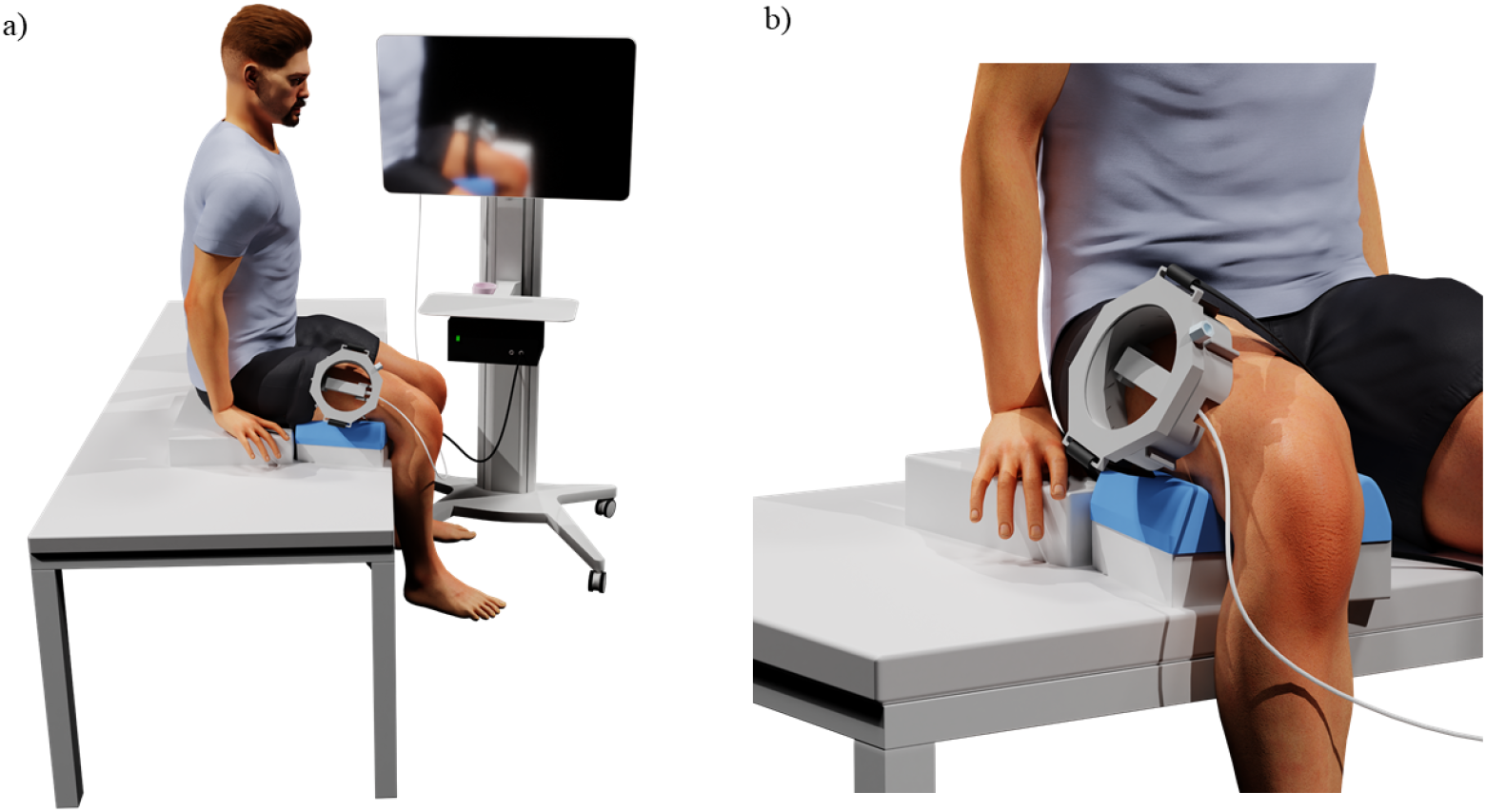
Measurement setup for performing THE on the VL muscle. (a) Participant seated upright with the right leg supported, hip at 90^*°*^ and knee at ≈90^*°*^ flexion. The ultrasound transducer was positioned at 75% of the distance between the greater trochanter and lateral femoral epicondyle, then secured with an elastic strap and a custom 3D-printed transducer holder allowing controlled rotations. (b) The holder enabled precise alignment of the transducer parallel and perpendicular to muscle fibers for longitudinal and transverse measurements.

Continuous multi-frequency mechanical vibrations (60, 70, 80 Hz) were induced by a custom designed pillow-driver (Elastance Imaging, Ohio, US). These drive frequencies were selected to yield shear wavelengths *λ* = *V*_*S*_*/f* in the ≈ 12–67 mm range for the expected *V*_*S*_ ≈ 1–4 m s^−1^ in vastus lateralis, matching muscle thickness and our field of view, while balancing attenuation (lower frequencies penetrate better) and ensuring sufficient energy delivery at depth and subject comfort. The ultrasound system acquired radio-frequency (RF) data line-by-line at 100 frames s^−1^ for 12 s per acquistion. Given the 100 Hz frame rate (Nyquist frequency 50 Hz), the 60/70/80 Hz inputs alias to 40/30/20 Hz in the recorded time series; we account for this with controlled-aliasing reconstruction to recover the true frequency components [27].

Three contraction states were tested in each orientation: (i) **rest** (no external load), (ii) **15% MVC**, and (iii) **30% MVC**. MVC was determined in a preliminary trial with the participant seated on a custom-built chair and the lower leg individually adjusted and attached to a load cell (SM-2000N, Interface Inc., Scottsdale) with a strap secured above the ankle joint. For each participant, this yielded six acquisitions (two orientations × three contraction levels); each condition comprised a continuous 12 s THE acquisition from which mean SWS was obtained by temporal and spatial averaging within the region of interest (ROI).

### 2.3 RF data processing

The THE pipeline followed Aghamiry et al. [26] and was applied identically to both transducer orientations to avoid processing-induced bias. For completeness, we summarize each step here and point to detailed derivations and design choices in the Appendices.

1. **Monogenic, phase-based displacement estimation:** Axial (vertical) tissue displacements between consecutive RF frames were estimated with a monogenic, phase-constancy optical-flow solver. The RF (or IQ-envelope) data were analyzed in a multiscale band-pass pyramid; at each scale, the monogenic signal (Riesz transform pair plus even component) yielded local amplitude, phase, and orientation in a rotation-invariant representation [28, 29]. Assuming small interframe motion, phase constancy was linearized to relate temporal and spatial monogenic phase gradients to the vertical displacement. We solved the resulting weighted least-squares problem with Tikhonov regularization via Gauss–Newton updates on a coarse-to-fine pyramid (see Appendices A and B for full formulae). To improve SNR while preserving coherent wave structure, we applied truncated-SVD denoising to the space–time stack, retaining eigenimages of ranks 2–100 [25, 30].
2. **Harmonic isolation:** Because THE used continuous multi-frequency harmonic driving, we isolated each vibration frequency with third-order Butterworth band-pass filters (center 60/70/80 Hz;*±*1.5 Hz bandwidth), yielding complex, frequency- and time-resolved displacement fields for each vibration frequency.
3. **Directional filtering:** Our ultrasound acquisition measured predominantly the vertical displacement component. For shear waves that propagate horizontally in-plane, particle motion is largely vertical (shear–vertical, SV-like), which maximizes sensitivity of the measured component and stabilizes the inversion. Conversely, oblique/vertical propagation introduces (i) mode mixing (SV/SH and guided components; SH denotes shear–horizontal, i.e., particle motion perpendicular to the vertical plane containing the propagation direction), (ii) stronger boundary/reflection contamination, and (iii) spatial interference that biases wavelength estimates. We therefore applied a purely angular Gaussian filter in the 2-D wavenumber domain to retain horizontally propagating energy while leaving the radial wavenumber |**k**| (and thus dispersion) unchanged [31, 32]. Specifically, we discretized propagation angle into *N* =32 directions and summed two antipodal windows centered at 0^*°*^ and 180^*°*^ with angular standard deviation *σ* = 2*π/N* = *π/*16 (FWHM ≈26.5^*°*^); see Fig. 2 for the filter shape and Appendix C for definitions.
4. **Wavenumber-based inversion (***k***-MDEV):** Wave number (k) based multifrequency dual elasto visco (k-MDEV) inversion computes the local phase gradient to estimate the wavenumber. These are averaged across directional components and vibration frequencies. Each timepoint of the time-resolved wavefields is processed individually, yielding a time-resolved SWS maps with 1 mm in-plane resolution.

**Fig. 2.**
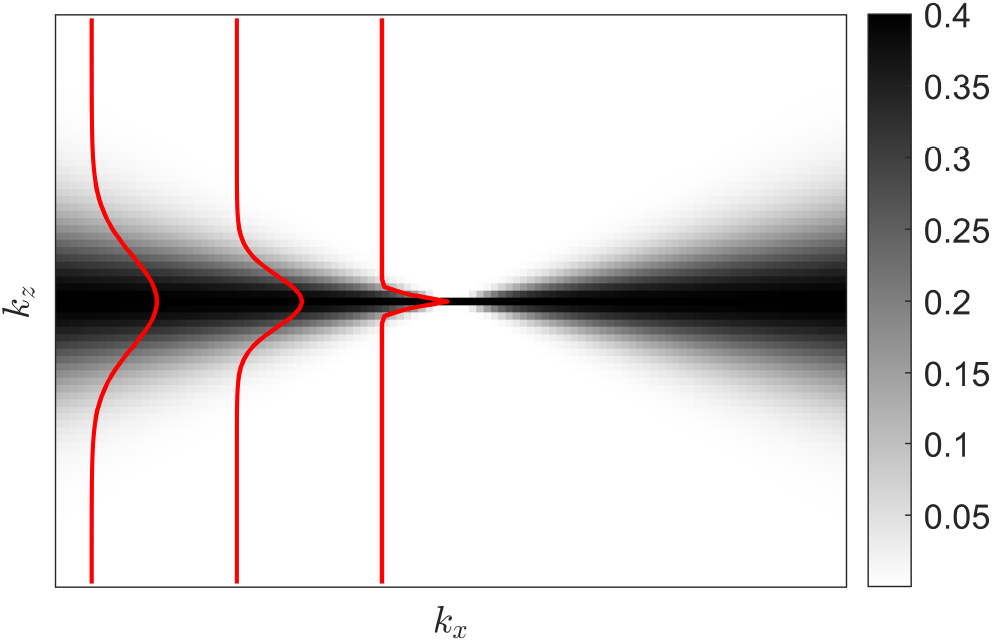
Directional Gaussian filter in the (*k*_*x*_, *k*_*z*_) domain. Two angular Gaussian windows (*N* = 32, *σ* = *π/*16) centered at 0^*°*^ and 180^*°*^ were summed to pass horizontally propagating components while attenuating others (refer to Appendix C for more details). Selected columns of the 2-D filter are highlighted in red to visualize angular bandwidth at different *k*_*x*_.

For each measurement, a region of interest (ROI) was drawn on the first B-mode frame to include VL fascicles. To account for in-plane motion, we performed frame-to-frame rigid registration by cross-correlation and propagated the resulting translation to the ROI across all 1,200 frames (12 s). Mean SWS values for the two transducer orientations were then obtained by temporal+spatial averaging within the ROI: *SWS*_∥_ for longitudinal scans (transducer aligned with fibers) and *SWS*_⊥_ for transverse scans (transducer perpendicular to fibers).

### 2.4 Pennation–angle correction

In pennate muscle, the transducer’s lateral imaging axis (dominant propagation direction in our setup) was rarely perfectly aligned with fascicles. A small in-plane fascicle angle *θ*_pen_ therefore mixed along- and across-fiber shear responses, biasing the apparent SWS. Because muscle was well approximated as a TI medium with symmetry along the fibers, we corrected this bias by mapping the apparent speeds measured in the transducer frame 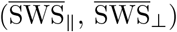 to the principal (fiber) frame (SWS_∥_, SWS_⊥_). Under small-strain, quasi-incompressible TI assumptions, the in-plane shear mode follows a directional harmonic-mean relation; solving the paired inverse problem yielded (see Appendix D):

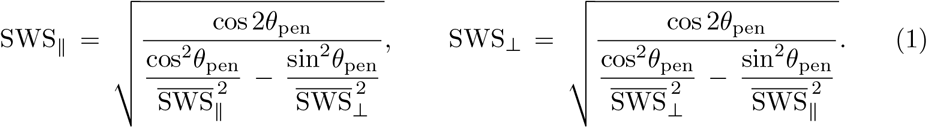

Fig. 3 schematizes this mapping: the transducer–frame measurements 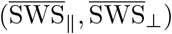 are projections of the fibre–frame ellipse rotated by *θ*_pen_, and Eq. (1) back-rotates them to (SWS_∥_, SWS_⊥_).

**Fig. 3.**
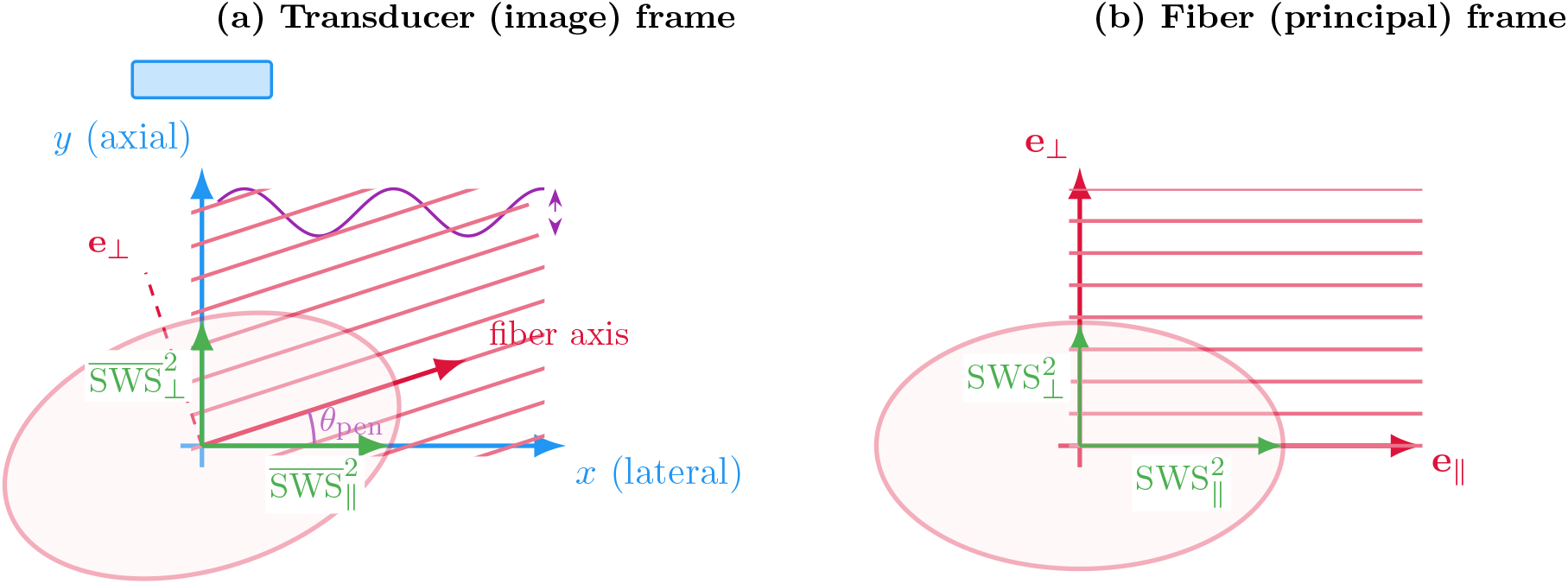
Schematic of coordinate frames and fascicle angle for pennation correction. **(a)** transducer (image) frame: the imaging plane was rotated by *θ*_pen_ relative to the fibers (purple stripes). The blue rectangle at the top represents the ultrasound transducer, centered on the *y*-axis. A harmonic wave was drawn below the transducer to illustrate a shear wave propagating horizontally (along +*x*). The vertical arrow on the *y*-axis indicated the direction of displacement estimation from RF phase. Green arrows mark the apparent squared speeds 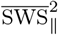 (lateral) and 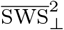 (axial). **(b)** Fiber (principal) frame: Eq. (1) maps the apparent measurements to SWS_∥_ and SWS_⊥_.

We applied Eq. (1) to each measurement using the frame-wise *θ*_pen_ estimated from the B-mode (below). As reported by Lima et al. [2], Miyamoto et al. [16], because |*θ*_pen_| ≲ 20^*°*^ in VL, corrections were small but systematic and avoided bias in *AI*.

#### Estimating θ_pen_ from B-mode

Angles were extracted directly from B-mode; the procedure, based on monogenic orientation (Appendix A), is summarized below:

1. *Monogenic features and phase asymmetry:* For each B-mode frame we computed a multiscale monogenic signal, yielding local orientation (Fig. 4b) and a phase-asymmetry (PA) map (Fig. 4a) that highlights line-like fascicular texture.
2. *Fiber mask:* Within the ROI, a PA threshold (*T* =0.1) selected fascicle-dominated pixels, suppressing non-fascicular edges.
3. *Orientation sampling and histogramming:* Monogenic orientations were sampled only inside the fiber mask, wrapped to [−90^*°*^, 90^*°*^], and histogrammed at high angular resolution.
4. *Gaussian fit and frame angle:* A single Gaussian *a* exp[−(*α* − *µ*)^2^*/σ*^2^] was fitted to the histogram; the fitted mean *µ* gave the frame-wise *θ*_pen_. Frames with insufficient masked pixels or poor fit (*R*^2^ *<* 0.9) were excluded. Per trial, the median of valid frame angles was used (5-frame median smoothing).

**Fig. 4.**
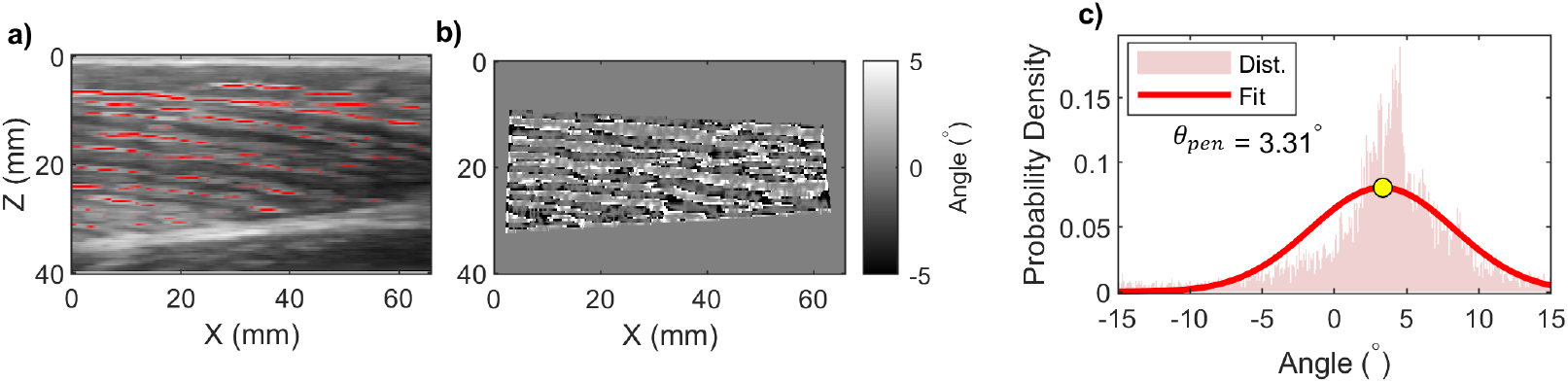
Pennation-angle estimation from a B-mode frame. **(a)** B-mode with monogenic phase-asymmetry overlay (red) highlighting fascicles. **(b)** Monogenic orientation (ROI). **(c)** Orientation histogram of masked pixels with Gaussian fit (red) and peak angle *θ*_pen_ (yellow).

The resulting *θ*_pen_ was then used in Eq. (1) to obtain (SWS_∥_, SWS_⊥_) from 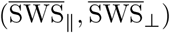 and to compute the corrected *AI* = SWS_∥_*/*SWS_⊥_. Figure 4 illustrates the angle-extraction steps.

### 2.5 Statistical analysis

Unless otherwise noted, all tests were two-sided with *α* = 0.01; results are reported as *p <* 0.01. Descriptive statistics are mean *±* SD.

To test how SWS depended on contraction level and transducer orientation, we used a two-factor repeated-measures ANOVA with intra-individual factors *Contraction* (0%, 15%, 30% MVC) and *Orientation* (longitudinal, transverse), evaluating main effects and their interaction. For AI, we used a one-factor repeated-measures ANOVA with intra-individual factor *Contraction* (0%, 15%, 30% MVC). Assumptions were checked on model residuals: normality with Shapiro–Wilk tests and Q–Q plots; sphericity with Mauchly’s test, applying Greenhouse–Geisser corrections when violated. With two levels, sphericity did not apply to *Orientation*. For the three-level *Contraction* factor, Bonferroni-adjusted post hoc pairwise comparisons were conducted where appropriate.

For intra-individual pairwise contrasts, Bonferroni-adjusted post hoc paired *t*-tests were conducted within each comparison family. For larger families of exploratory correlations, we controlled the false discovery rate (FDR) using the Benjamini–Hochberg procedure (*q* = 0.05). Effect sizes were reported as partial 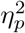 for ANOVAs and Cohen’s *d*_*z*_ for paired contrasts; 95% confidence intervals for *d*_*z*_ were obtained by percentile bootstrap (5,000 resamples).

To summarize contraction dependence, we computed per-subject linear slopes of SWS (and AI) versus MVC level (0, 15, 30%). For each orientation, the mean slope was tested against zero (one-sample *t*-test), and slopes were compared between orientations using a paired *t*-test.

Potential influences of participant characteristics were evaluated via Pearson correlations (age, mass, height, BMI, MVC) with FDR control across outcomes. We also screened for sex differences using independent-samples comparisons at each contraction level (FDR across tests).

## 3 Results

### 3.1 Pennation–angle correction

Measurements were successful in all 26 participants in both transducer orientations and MVC levels. SWS increased with contraction in both orientations, with a larger increase along fibers, however, these values must be adjust to account for the pennation angle. Using the monogenic orientation pipeline, in–plane fascicle angles were small at all loads (|*θ*_pen_| ≲ 10^*°*^). Signed means (SD) were: rest 1.9^*°*^ (2.7^*°*^), 15% MVC −0.8^*°*^ (3.2^*°*^), and 30% MVC 1.5^*°*^ (3.6^*°*^).

Applying Eq. 1 to SWS, produced small increases in anisotropy while preserving the contraction–dependent pattern: group–mean Δ*AI* = *AI*^corr^ −*AI*^meas^ were *∼*0.0001 (rest), *∼*0.0002 (15% MVC), and *∼*0.0003 (30% MVC). Paired *t*-tests were statistically significant at all loads (rest and 30%: *p <* 0.01; 15%: *p <* 0.01), but the absolute changes were negligible at the group level and did not alter interpretation. Individual volunteers exhibited noticeable paired shifts (Fig. 5), motivating retention of the correction for subject–level analyses.

**Fig. 5.**
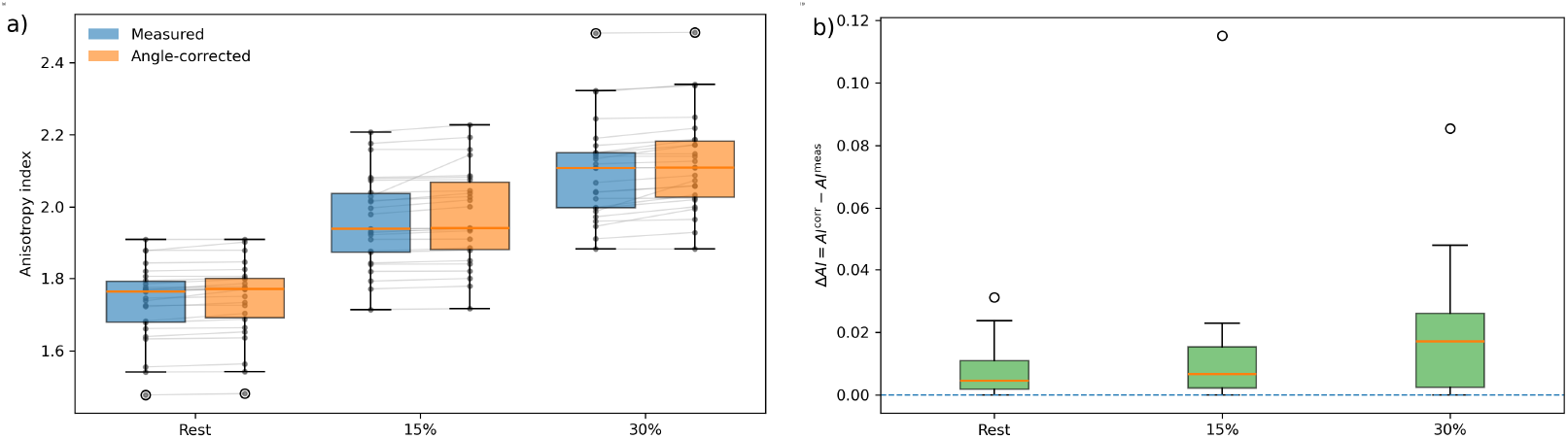
Effect of pennation correction (Eq. (1)) on anisotropy. **(a)** Box plots of *AI* at rest, 15% and 30% MVC for measured (blue) and angle-corrected (orange) data; gray lines connect each participant to visualize intra-individual shifts. **(b)** Box plots of Δ*AI* = *AI*^corr^ −*AI*^meas^. Corrections are negligible at the group level.

### 3.2 SWS maps in a representative volunteer

Figure 6 illustrates the angle dependence and spatial distribution of SWS in a representative volunteer at rest, 15% MVC, and 30% MVC. The first column shows polar plots of time-averaged SWS obtained with the directional filter bank (same as Fig. 2) rotated in 11.25^*°*^ steps. Longitudinal acquisitions are shown in blue and transverse acquisitions in red for different loads (rest, 15% MVC, and 30% MVC) in panels (a), (d), and (g), respectively. Across all angles, transverse SWS was lower than longitudinal SWS. In this study, we focused on waves that propagate horizontally in-plane (particle motion predominantly vertical), as these maximize sensitivity of the measured displacement component and stabilize the inversion.

**Fig. 6.**
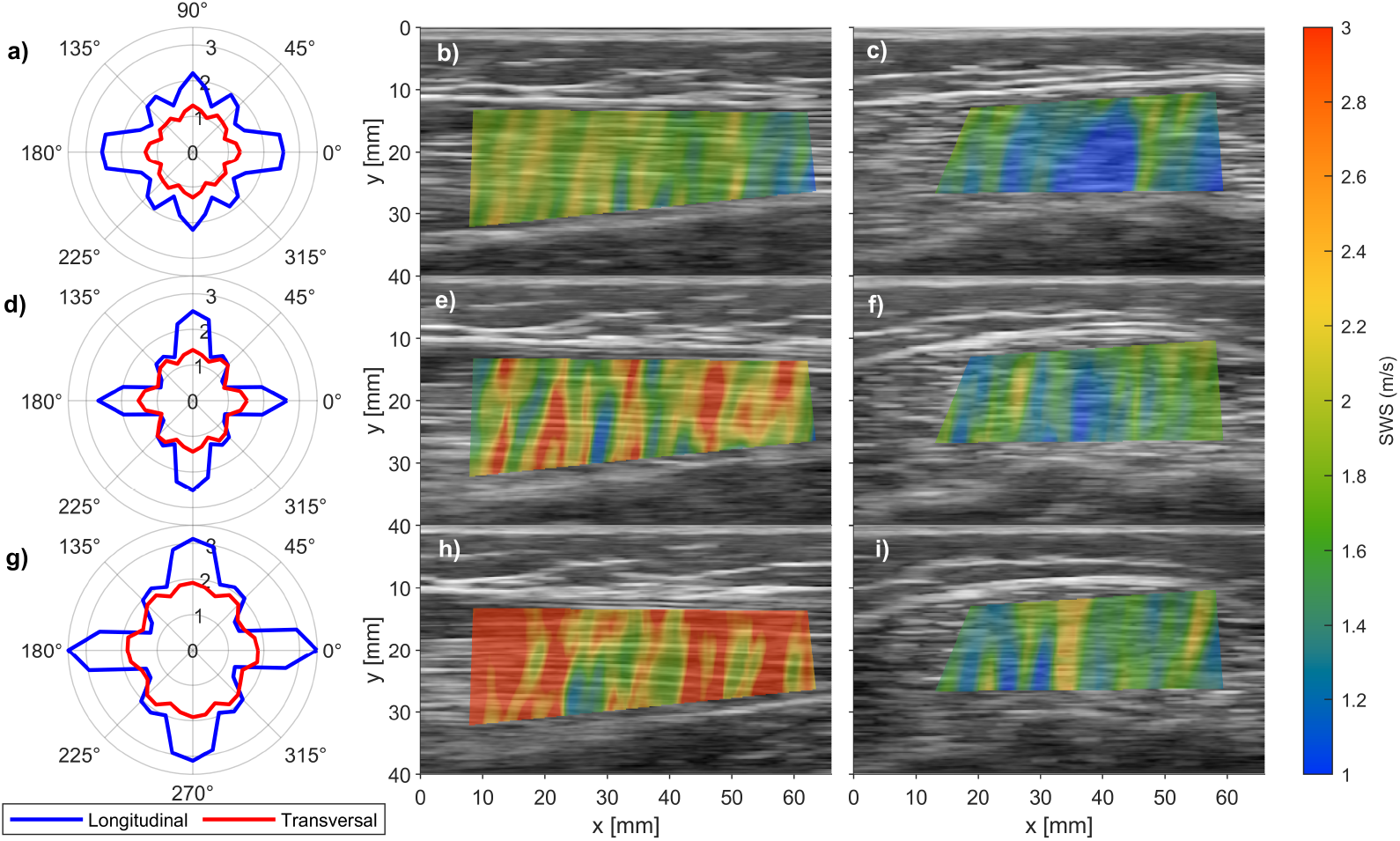
Representative example of angle-dependent and spatial SWS changes during isometric contraction. **(a**,**d**,**g)** Polar plots of mean SWS within the ROI using directional filters rotated in 11.25^*°*^ increments (same filter bank as Fig. 2). Longitudinal (blue) and transverse (red) measurements are shown for rest, 15% MVC, and 30% MVC. **(b**,**e**,**h)** Longitudinal SWS maps reconstructed from horizontally propagating waves and averaged over 12 s for rest, 15% MVC, and 30% MVC. **(c**,**f**,**i)** Corresponding transverse SWS maps. Underlying B-mode images are shown for anatomical reference. With increasing MVC, SWS increases in both orientations but less across fibers, yielding a higher *AI*.

The second and third columns display SWS maps reconstructed from horizontally propagating waves and averaged over 12 s within the selected ROI. Panels (b), (e), and (h) show longitudinal SWS maps for different loads; panels (c), (f), and (i) show the corresponding transverse maps. SWS increased from rest to 15% and 30% MVC in both orientations, with a smaller rate of increase across fibers. Consequently, the anisotropy index increased with contraction level.

### 3.3 Statistical analysis

As it was mentioned before, SWS increased with contraction in both orientations, with a larger increase along fibers. Group means (mean±SD) were: longitudinal SWS 2.5 *±* 0.2 (rest), 3.2 *±* 0.2 (15% MVC), 3.8 *±* 0.3 m/s (30% MVC); transverse SWS 1.4 *±* 0.1, 1.6 *±* 0.1, 1.8 *±* 0.1 m/s; and *AI*: 1.7 *±* 0.1, 2.0 *±* 0.1, 2.1 *±* 0.1, respectively. Box plots for all volunteers are shown in Fig. 7; gray lines connect individual participants, illustrating intra-individual increases from rest to higher MVC. The transverse increase was smaller than the longitudinal increase, yielding a rising *AI* across contraction levels.

**Fig. 7.**
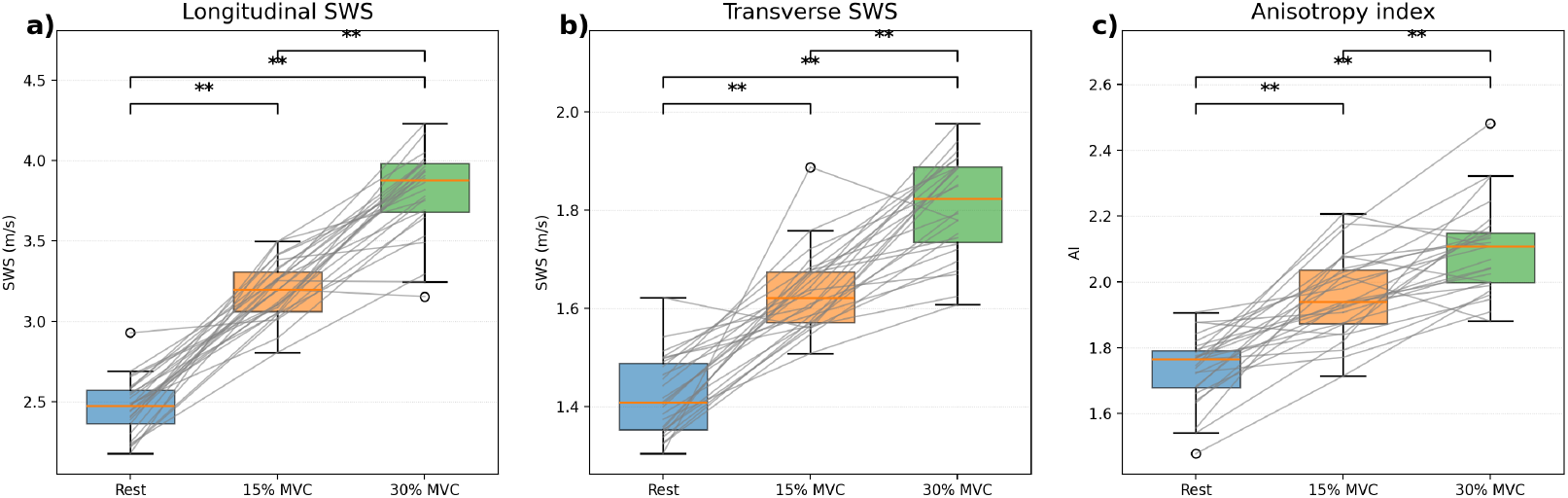
Panels (a–c) show longitudinal SWS, transverse SWS, and AI at 0% (rest), 15%, and 30% MVC. Colored box plots depict the distribution across participants and gray lines connect each participant across contraction levels to visualize intra-individual changes. Brackets and asterisks denote Bonferroni-adjusted post-hoc paired comparisons (** *p <* 0.01).

A two-factor repeated-measures ANOVA on SWS showed strong main effects of Orientation 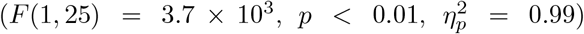 and contraction 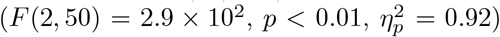, and a significant orientation*×* Contraction interaction 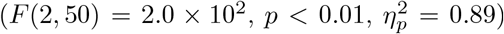. For *AI*, the one-factor RM-ANOVA was also significant 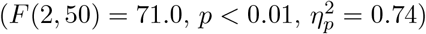.

A priori intra-individual contrasts, aligned with our hypotheses, compared changes across contraction levels (0→15, 15→30, 0→30% MVC) within each orientation and longitudinal vs. transverse at each level. These contrasts were tested with two-sided paired *t*-tests and Bonferroni adjustment within each comparison family (*p <* 0.01). Results are summarized in Table 2. Briefly, longitudinal SWS increased by +0.73 and +0.60 m/s from 0→15 and 15→30% MVC, respectively; transverse SWS increased by +0.21 and +0.18 m/s. *AI* increased by +0.22 (0→15%) and +0.14 (15→30%), and the difference between longitudinal and transverse SWS widened from +1.04 m/s at rest to +1.99 m/s at 30% MVC (all *p <* 0.01).

**Table 2.** Planned (pre-specified) pairwise contrasts. Entries are mean intra-individual differences (Δ, mean±SD), Cohen’s *d*_*z*_, and Bonferroni-adjusted *p* thresholds.

| Measure | Contrast | Mean $\Delta$ (±SD) | $d_z$ | Adj. $p$ |
| --- | --- | --- | --- | --- |
| Longitudinal SWS | 0→15 | $0.7 \pm 0.3$ | 2.8 | $< 0.01$ |
| Longitudinal SWS | 15→30 | $0.6 \pm 0.3$ | 2.1 | $< 0.01$ |
| Longitudinal SWS | 0→30 | $1.3 \pm 0.3$ | 4.6 | $< 0.01$ |
| Transverse SWS | 0→15 | $0.2 \pm 0.1$ | 1.8 | $< 0.01$ |
| Transverse SWS | 15→30 | $0.2 \pm 0.1$ | 1.6 | $< 0.01$ |
| Transverse SWS | 0→30 | $0.4 \pm 0.1$ | 3.0 | $< 0.01$ |
| AI | 0→15 | $0.2 \pm 0.2$ | 1.5 | $< 0.01$ |
| AI | 15→30 | $0.1 \pm 0.1$ | 1.0 | $< 0.01$ |
| AI | 0→30 | $0.4 \pm 0.2$ | 2.1 | $< 0.01$ |
| L vs T | Rest (L–T) | $1.0 \pm 0.1$ | 7.2 | $< 0.01$ |
| L vs T | 15% (L–T) | $1.6 \pm 0.2$ | 8.9 | $< 0.01$ |
| L vs T | 30% (L–T) | $2.0 \pm 0.2$ | 8.6 | $< 0.01$ |

Expressed as relative change from 0→30% MVC, longitudinal SWS increased by 54.0% and transverse SWS by 27.2%; *AI* increased by 21.1%. Individual linear trend slopes were positive for both orientations (longitudinal: 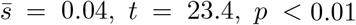; transverse: 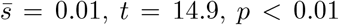) and were larger longitudinally than transversely (mean difference 0.03, *t* = 18.8, *p <* 0.01, *d*_*z*_ = 3.7). *AI* slopes were also positive 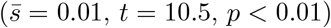. Finally, no sex differences or correlations with age, anthropometrics, or BMI survived FDR control across outcomes.

## 4 Discussion

Using THE, we quantified direction-dependent SWS in the VL muscle at rest and during low-level isometric contractions. Three results stand out. First, VL exhibited marked baseline anisotropy: longitudinal SWS exceeded transverse SWS at rest (2.5 *±* 0.2 vs. 1.4 *±* 0.1 m/s), yielding *AI* = 1.7 *±* 0.1. Second, SWS increased with activation in both orientations, but more along fibers (rest*→*30% MVC: +54.0% longitudinal, +27.2% transverse), producing a statistically significant rise in *AI* to 2.1*±* 0.1 (+21.1%). Third, a small in-plane pennation-angle correction (median |*θ*_pen_| ≲ 10^*°*^) produced small group-level shifts in anisotropy, leaving the contraction-dependent pattern unchanged. While negligible on average, subject-level shifts were sometimes noticeable (cf. Fig. 5a), supporting retention of the correction in the analysis pipeline.

Broadly, our orientation-specific SWS and the observed anisotropy are consistent with prior elastography work showing that skeletal muscle behaves as a transversely isotropic, direction-dependent medium [1, 3, 19, 21, 24]. Studies using single-orientation ultrasound (e.g., supersonic shear imaging (SSI)) and TI-aware reconstructions alike report higher along-fiber than across-fiber speeds at rest, with magnitudes depending on muscle, frequency, and wave-mode conditioning [2, 16–18].

Our findings in VL at rest closely match those obtained with rotational 3D SWEI by Knight et al. [17]. Using an ARFI-based 3D acquisition with transducer rotation and intentional fiber tilt, they reported shear moduli of *µ*_∥_ = 5.8 *±* 1.0 kPa and *µ*_⊥_ = 1.9*±*0.4 kPa in relaxed VL, corresponding to a shear anisotropy 𝒳_*µ*_ = (*µ*_∥_ −*µ*_⊥_)*/µ*_⊥_ = 2.1 *±* 0.9 and a tensile anisotropy 𝒳_*E*_ = 4.7 *±* 1.4. In the same dataset, representative group speeds were *c*_∥_ ≈ 2.3 m/s and *c*_⊥_ ≈ 1.4 m/s (from ellipse fits to the SH mode). In our THE data, the corresponding relaxed values were SWS_∥_ = 2.5 *±* 0.2 m/s and SWS_⊥_ = 1.4 *±* 0.1 m/s, giving *AI* = 1.7 *±* 0.1. This is numerically close to the 3D SWEI speed ratio (*∼*1.7) and, via 𝒳_*µ*_ = *AI*^2^ ™ 1, implies a shear-modulus anisotropy of ≈1.9 at rest—again consistent with Knight et al. [17]. The agreement is encouraging given substantial protocol differences: Knight et al. [17] used ARFI pushes with rotational sampling to resolve both shear-horizontal (SH) and shear-vertical (SV) modes and to estimate three TI parameters (*µ*_∥_, *µ*_⊥_, 𝒳_*E*_); our THE approach uses continuous external vibrations (60–80 Hz), 2-D imaging, and directional filtering to isolate horizontally propagating waves, and we summarize anisotropy by the speed ratio *AI*.

Methodologically, these studies are complementary. The rotational 3D SWEI geometry leverages natural fiber tilt to excite and observe both SH and SV modes, enabling recovery of tensile anisotropy 𝒳_*E*_ and demonstrating that accounting for fiber tilt increases *µ*_∥_ estimates relative to 2-D methods. Our pipeline standardizes transducer placement, applies narrow-band angular filtering, and performs a small-angle (pennation) correction; while we do not recover 𝒳_*E*_, we obtain stable, field-of-view–wide SWS maps that reproduce the same baseline anisotropy in VL.

The greater activation-induced rise of SWS along fibers is consistent with tension-dependent shear behavior of actively cross-bridged myofibrils and with collagenous alignment that preferentially stiffens the fiber axis under load [4–6]. The persistence and amplification of anisotropy with low-level activation suggest that *AI* captures aspects of the force-bearing architecture beyond scalar stiffness. Whether *AI* saturates or changes nonlinearly at higher forces remains an open question.

Several limitations merit consideration. First, the external vibration source generates a superposition of wave modes and propagation directions; although angular filtering emphasizes in-plane, horizontally propagating shear waves, complete mode isolation is not guaranteed within the finite aperture [31]. Second, we measured only the vertical displacement component; in TI media both propagation direction and polarization govern phase velocity, which limits full tensor recovery of the third free elasticity-related parameter in an incompressible TI material, and may leave residual mixing. Third, the study covered a narrow force and frequency range (0–30% MVC, 60–80 Hz) in a healthy young cohort. Therefore, anisotropy values may depend on activation level, dispersion, or muscle condition. These factors must be taken into account when interpreting our data, since our results represent apparent direction-dependent SWS under a controlled but incomplete observation model.

Despite these limitations, the study shows that THE, combined with directional filtering and TI-aware interpretation, yields robust, orientation-sensitive metrics with tight confidence intervals and strong repeated-measures effects. Reporting SWS_∥_, SWS_⊥_, and *AI* avoids questionable isotropic conversions (*E ≈* 3*µ*) and is better tailored to muscle’s TI behavior [19, 21, 24]. The approach is simple to deploy at the bedside, requires no ARFI pulses or ultrafast plane-wave hardware, and produces consistent maps across contraction states, supporting applications in physiology (activation mapping, fatigue, training adaptation) and disease (e.g., dystrophies, spasticity) [13, 14, 22].

Two avenues are promising for future work. First, *measurement completeness*: capturing multi-component data (2.5D or 3D) and incorporating multi-component motion estimation enhances mode control and enables more comprehensive recovery of TI parameters. [3, 17, 18, 33]. Second, *biophysical integration*: combining THE with diffusion tensor imaging or fascicle-tracking ultrasound could link *AI* to microstructure, pennation dynamics, and force production [21, 34, 35]. Extending the frequency band and MVC range will clarify dispersion and force dependence; clinical studies can test whether direction-resolved metrics add diagnostic or prognostic value [15, 23, 36].

In sum, THE reveals a pronounced and activation-sensitive anisotropy of VL shear-wave propagation. Direction-resolved SWS and their ratio provide compact, reproducible biomarkers of muscle mechanical behavior that complement—and in some scenarios may outperform—isotropic elastography surrogates.

## 5 Conclusion

THE is a cost-effective ultrasound elastography approach that captures the intrinsic anisotropy of skeletal muscle. In this study, we applied THE for the first time to recover SWS longitudinal and transverse to the fiber direction in VL under different loading conditions in healthy volunteers. The measurements revealed marked tissue anisotropy that increased with contraction intensity. These direction-resolved SWS maps demonstrate the feasibility of using THE to characterize muscle mechanics in vivo and provide an orientation-sensitive readout aligned with muscle architecture particularly for low-level isometric contractions of knee extensor muscles. The reported SWS values establish a reference for THE in skeletal muscle and support future translation to clinical assessments. Together, these findings position THE as a practical tool for evaluating neuromuscular function with sensitivity to fiber direction.

## Data Availability

All data produced in the present study are available upon reasonable request to the authors

## Acknowledgments

This study was funded by AIF FKZ KK5611902 BM4 (MUSKEL), German Research Foundation (DFG) projects 513752256 FOR5628, CRC1340, GRK2260 BIOQIC, and GU 172614-1. These sponsors had no role in the study design, collection, analysis and interpretation of data, writing of this manuscript or the decision to submit the article for publication.

## Appendix A Monogenic signal and phase asymmetry

The monogenic signal was used as a 2-D generalization of the 1-D analytic signal to separate a band-limited image into an even (cosine-like) component and two odd (sine-like) quadrature components given by the Riesz transform [37–39]. This representation yielded three local, rotation-invariant descriptors (amplitude, phase, and orientation) that were well suited to ultrasound, where intensity could vary but geometry (phase) was relatively stable.

Let *f* (**x**, *t*) denote an image at time/frame *t* with **x** = (*x, y*). At scale *s* we computed a band-passed image

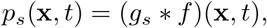

where *g*_*s*_ was an (approximately) isotropic band-pass filter (e.g., Gaussian–derivative or Riesz–Laplace wavelet). The 2-D Riesz transform ℛ= *R*_1_, *R*_2_ acted through its Fourier symbol

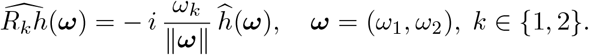

The monogenic triplet at scale *s* was

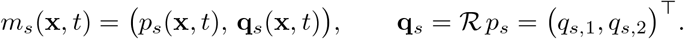

From (*p*_*s*_, **q**_*s*_) we obtained

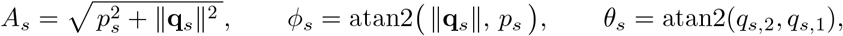

i.e., local amplitude *A*_*s*_, local phase *ϕ*_*s*_, and orientation angle *θ*_*s*_ (direction of maximal variation). The phase *ϕ*_*s*_ is known to be contrast-invariant, which improves robustness for ultrasound.

### Phase asymmetry (PA) for fiber detection

To emphasize line-like texture (muscle fascicles and aponeuroses), we used a phase-based feature known as phase asymmetry (PA) [40]. With the scale-consistent phase *ϕ*_*s*_ we defined

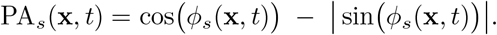

PA was strongly positive on ridges/lines (central bright structure), provided a characteristic negative–positive transition across step edges, and was relatively insensitive to intensity scaling. In our workflow, PA was computed on the B-mode (envelope of the RF/IQ data) and thresholded within the ROI to produce a fiber mask that suppressed non-fascicular texture. Monogenic orientations *θ*_*s*_ were then sampled only from masked pixels and aggregated across scales to form a high-resolution orientation histogram used to estimate the frame-wise fascicle angle.

## Appendix B Monogenic displacement estimation method

We estimated the axial (vertical) displacement between consecutive RF frames using a monogenic, phase-constancy optical-flow formulation [28, 29, 41–43]. The key assumption, valid for small interframe motion, was that local phase was conserved along motion trajectories. Specializing the general phase-constancy linearization to the vertical direction *y* gave

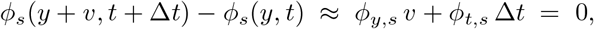

so the per-scale vertical displacement per frame was

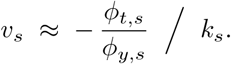

Here *k*_*s*_ converted phase (radians) to cycles/pixel for the chosen band-pass (for a Gaussian-derivative band-pass with scale *s, k*_*s*_ *≈ ω*_0_(*s*)*/*(2*π*) with *ω*_0_ ∝ *s*^−1*/*2^). A stable estimate of the vertical phase derivative was written using the monogenic components [28, 29]:

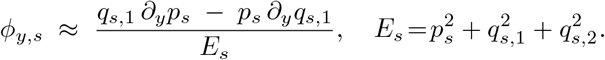

Between two consecutive frames, a rotation-invariant signed phase difference was obtained from inner and cross products of monogenic parts:

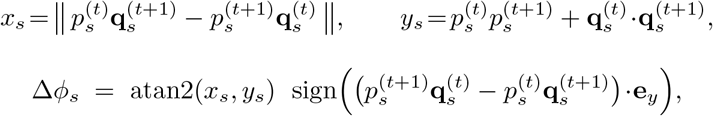

with **e**_*y*_ the vertical unit vector.

Aggregating scales 𝒮, we estimated the vertical displacement field *v* by solving a weighted least-squares problem. More stable estimates were achieved by adding Tikhonov regularization:

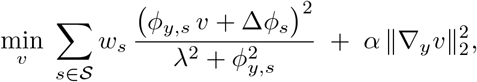

where *w*_*s*_ upweighted reliable scales (e.g., proportional to *E*_*s*_), *λ* stabilized low-gradient regions, and *α* was the smoothness weight. We solved this with Gauss–Newton updates on a coarse-to-fine pyramid and frame-to-frame warping to converge for larger motions.

In 1-D without band-pass filtering and regularization, this approach reduced to classic phase-shift estimation related to Hilbert/analytic-signal methods [44].

## Appendix C Directional angular filtering in the wavenumber domain

External vibration excited a mixture of wave modes and directions. Because we measured only the vertical displacement component (as most ultrasound methods do), we emphasized in-plane shear waves that propagated horizontally, whose particle motion was largely vertical (SV-like). To this end we used a purely angular Gaussian filter in the 2-D wavenumber domain (*k*_*x*_, *k*_*z*_), which preserved the radial wavenumber 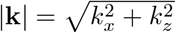 (and thus dispersion) while selecting propagation direction [31].

Let

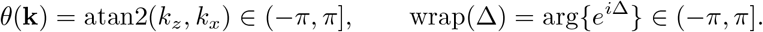

We discretized the unit circle into *N* = 32 directions

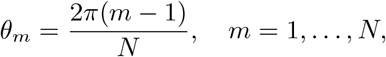

and selected the antipodal horizontal directions *I* = {1, 17} corresponding to 0^*°*^ and 180^*°*^. For each selected direction we defined a Gaussian angular window

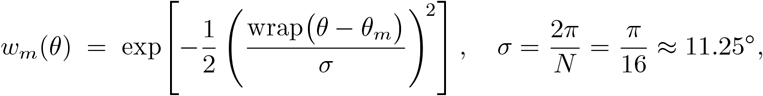

and set the directional passband as the sum

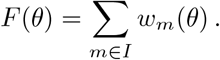

The full width at half maximum (FWHM) of each lobe was 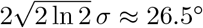. We set *F* (*θ*) = 0 at **k** = **0** (undefined angle). This filter bank passed horizontally propagating energy in both ±*k*_*x*_ directions and attenuated oblique or vertical propagation that was more susceptible to boundary reflections, mode conversion, and standing-wave interference.

## Appendix D Derivation of the pennation-angle correction

Skeletal muscle was modeled as transversely isotropic (TI) with the symmetry axis aligned to fibers. When the transducer’s lateral imaging axis was rotated by an in-plane angle *θ* relative to fibers, the measured speeds 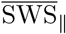 (lateral) and 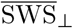 (axial) were apparent values that mixed the true along- and across-fiber shear responses. Under small strain and quasi-incompressibility, the effective in-plane shear modulus followed a directional harmonic-mean relation [33]:

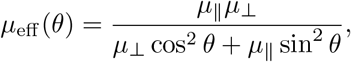

where *µ*_∥_ was the (higher) shear modulus for propagation exactly along fibers and *µ*_⊥_ was for propagation exactly across fibers.

Assuming constant density *ρ*, the shear-wave speed satisfied 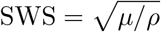. Hence the measured (apparent) speeds at angle *θ* and its orthogonal complement obeyed

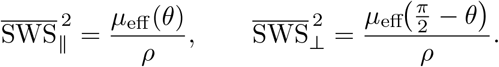

Let 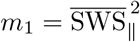 and 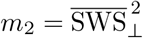. Using the harmonic-mean form above and writing *c* = cos *θ, s* = sin *θ*, we obtained a linear system in the inverse moduli *x* = 1*/µ*_∥_ and *y* = 1*/µ*_⊥_:

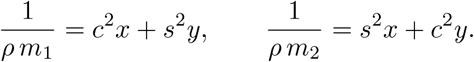

Solving using *c*^4^ − *s*^4^ = cos 2*θ* gave

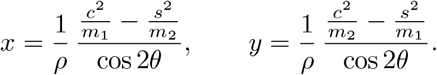

Therefore,

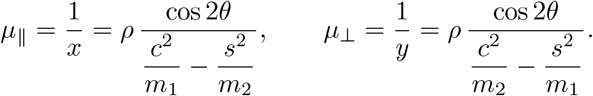

Finally, we converted back to the true along- and across-fiber speeds via 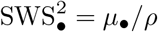, yielding

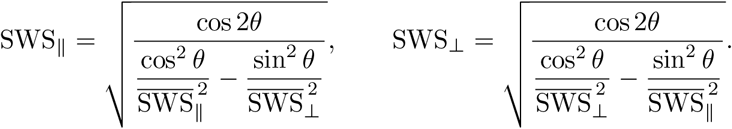

## References

[1] Gennisson, J.L., Catheline, S., Chaffaý, S., Fink, M.: Transient elastography in anisotropic medium: application to the measurement of slow and fast shear wave speeds in muscles. The Journal of the Acoustical Society of America 114(1), 536–541 (2003)

[2] Lima, K., Rouffaud, R., Grégoire, J.M., et al.: Supersonic shear imaging elastography in skeletal muscles: Relationship between in vivo and synthetic fiber angles and shear modulus. Journal of Ultrasound in Medicine 38(1), 81–90 (2019) 18

[3] Rouze, N.C., Palmeri, M.L., Nightingale, K.R.: Full characterization of in vivo muscle as an elastic, incompressible, transversely isotropic material using ultrasonic rotational 3d shear wave elasticity imaging. IEEE Transactions on Medical Imaging 41(1), 133–144 (2022)

[4] Takaza, M., Moerman, K.M., Gindre, J., Lyons, G., Simms, C.K.: The anisotropic mechanical behaviour of passive skeletal muscle tissue subjected to large tensile strain. Journal of the Mechanical Behavior of Biomedical Materials 17, 209–220 (2013)

[5] Maas, H., Sandercock, T.G.: Force transmission between synergistic skeletal muscles through connective tissue linkages. BioMed Research International 2010, 575672 (2010)

[6] Ramaswamy, K.S., Palmer, M.L., Meulen, J.H., Renoux, A., et al.: Lateral transmission of force is impaired in skeletal muscles of dystrophic mice and very old rats. The Journal of Physiology 589, 1195–1208 (2011)

[7] Nakamura, M., Ikezoe, T., Takeno, Y., Ichihashi, N.: Effects of a 4-week static stretch training program on passive stiffness of human gastrocnemius muscletendon unit in vivo. European Journal of Applied Physiology 112, 2749–2755 (2012)

[8] Seynnes, O.R., Boer, M., Narici, M.V.: Early skeletal muscle hypertrophy and architectural changes in response to high-intensity resistance training. Journal of Applied Physiology 102, 368–373 (2007)

[9] Preobrazenski, N., Seigel, J., Halliday, S., Janssen, I., McGlory, C.: Single-leg disuse decreases skeletal muscle strength, size, and power in uninjured adults: A systematic review and meta-analysis. Journal of Cachexia, Sarcopenia and Muscle 14, 684–696 (2023)

[10] Abboud, J., Lardon, A., Boivin, F., Dugas, C., Descarreaux, M.: Effects of muscle fatigue, creep, and musculoskeletal pain on neuromuscular responses to unexpected perturbation of the trunk: a systematic review. Frontiers in Human Neuroscience 10, 667 (2017)

[11] Catheline, S., Gennisson, J.-L., Delon, G., Fink, M., Sinkus, R., Abouelkaram, S., Culioli, J.: Measurement of viscoelastic properties of homogeneous soft solid using transient elastography: An inverse problem approach. The Journal of the Acoustical Society of America 116(6), 3734–3741 (2004)

[12] Deffieux, T., Gennisson, J.-L., Tanter, M., Fink, M.: Assessment of the mechanical properties of the musculoskeletal system using 2-d and 3-d very high frame rate ultrasound. IEEE transactions on ultrasonics, ferroelectrics, and frequency control 55(10), 2177–2190 (2008)

[13] Brandenburg, J.E., Eby, S.F., Song, P., Zhao, H., Brault, J.S., Chen, S., An, K.N.: Ultrasound elastography: the new frontier in direct measurement of muscle stiffness. Archives of Physical Medicine and Rehabilitation 95(11), 2207–2219 (2014)

[14] Ngo, H.H.P., Poulard, T., Brum, J., et al.: Anisotropy in ultrasound shear wave elastography: An add-on to muscles characterization. Frontiers in Physiology 13, 1000612 (2022)

[15] Yang, Y., Shahryari, M., Meyer, T., Garcia, S.R.M., Görner, S., Majd, M.S., Guo, J., Braun, J., Sack, I., Tzschätzsch, H.: Explorative study using ultrasound time-harmonic elastography for stiffness-based quantification of skeletal muscle function. Zeitschrift für Medizinische Physik (2024)

[16] Miyamoto, N., Hirata, K., Kanehisa, H., et al.: Validity of measurement of shear modulus by ultrasound shear wave elastography in human pennate muscle. PLOS One 10(4), 0124311 (2015)

[17] Knight, A.E., Trutna, C.A., Rouze, N.C., Hobson-Webb, L.D., Caenen, A., Jin, F.Q., Palmeri, M.L., Nightingale, K.R.: Full characterization of in vivo muscle as an elastic, incompressible, transversely isotropic material using ultrasonic rotational 3d shear wave elasticity imaging. IEEE transactions on medical imaging 41(1), 133–144 (2021)

[18] Wightman, W.E., Rouze, N.C., Chan, D.Y., Srinivasan, S., Palmeri, M.L., Nightingale, K.R.: Multidimensional swei algorithms in rotationally sampled media: Assessing the accuracy of phase velocity reconstruction in transversely isotropic media. In: 2024 IEEE Ultrasonics, Ferroelectrics, and Frequency Control Joint Symposium (UFFC-JS), pp. 1–4 (2024). IEEE

[19] Papazoglou, S., Rump, J., Braun, J., Sack, I.: Shear wave group velocity inversion in mr elastography of human skeletal muscle. Magnetic Resonance in Medicine 56(3), 489–497 (2006)

[20] Ma, S., He, Z., Wang, R., Zhang, A., Sun, Q., Liu, J., Yan, F., Sacks, M.S., Feng, X.-Q., Yang, G.-Z., et al.: Measurement of biomechanical properties of transversely isotropic biological tissue using traveling wave expansion. Medical Image Analysis 101, 103457 (2025)

[21] Guo, J., Hirsch, S., Scheel, M., Braun, J., Sack, I.: Three-parameter shear wave inversion in mr elastography of incompressible transverse isotropic media: Application to in vivo lower leg muscles. Magnetic Resonance in Medicine 75(4), 1537–1545 (2016)

[22] Moore, C.L., Gallardo, M.R., Sniadecki, N.J., et al.: In vivo viscoelastic response (visr) ultrasound for characterizing mechanical anisotropy in lower-limb skeletal muscles of boys with and without duchenne muscular dystrophy. Ultrasound in Medicine & Biology 44(12), 2519–2530 (2018)

[23] Ringleb, S.I., Bensamoun, S.F., Chen, Q., Manduca, A., An, K.-N., Ehman, R.L.: Applications of magnetic resonance elastography to healthy and pathologic skeletal muscle. Journal of Magnetic Resonance Imaging: An Official Journal of the International Society for Magnetic Resonance in Medicine 25(2), 301–309 (2007)

[24] Klatt, D., Papazoglou, S., Braun, J., Sack, I.: Viscoelasticity-based mr elastography of skeletal muscle. Physics in Medicine & Biology 55(21), 6445–6459 (2010)

[25] Meyer, T., Wellge, B., Barzen, G., Chandia, S.K., Knebel, F., Hahn, K., Elgeti, T., Fischer, T., Braun, J., Tzschätzsch, H., et al.: Cardiac elastography with external vibration for quantification of diastolic myocardial stiffness. Journal of the American Society of Echocardiography 38(5), 431–442 (2025)

[26] Aghamiry, H.S., Meyer, T., Klemmer Chandía, S., et al.: Muscle activation assessment using ultrasound time-harmonic elastography and tonic vibration reflex. Preprint (2025)

[27] Tzschätzsch, H., Guo, J., Dittmann, F., Hirsch, S., Barnhill, E., Jöhrens, K., Sack, I.: Two-dimensional time-harmonic elastography of the human liver and spleen. Ultrasound in Medicine Biology 42(11), 2562–2571 (2016)

[28] Felsberg, M.: Optical flow estimation from monogenic phase. In: Complex Motion (IWCM 2004). Lecture Notes in Computer Science, vol. 3417, pp. 1–13. Springer, ??? (2007)

[29] Unser, M., Sage, D., Van De Ville, D.: Multiresolution monogenic signal analysis using the riesz–laplace wavelet transform. IEEE Transactions on Image Processing 18(11), 2402–2418 (2009)

[30] Hansen, P.C.: The truncated svd as a method for regularization. BIT Numerical Mathematics 27(4), 534–553 (1987)

[31] Manduca, A., Lake, D.S., Kruse, S.A., Ehman, R.L.: Spatio-temporal directional filtering for improved inversion of mr elastography images. Medical image analysis 7(4), 465–473 (2003)

[32] Tzschätzsch, H., Guo, J., Dittmann, F., Hirsch, S., Barnhill, E., Jöhrens, K., Braun, J., Sack, I.: Tomoelastography by multifrequency wave number recovery from time-harmonic propagating shear waves. Medical image analysis 30, 1–10 (2016)

[33] Wang, M., Byram, B., Palmeri, M., Rouze, N., Nightingale, K.: Imaging transverse isotropic properties of muscle by monitoring acoustic radiation force induced shear waves using a 2-d matrix ultrasound array. IEEE transactions on medical imaging 32(9), 1671–1684 (2013)

[34] Heemskerk, A.M., Strijkers, G.J., Vilanova, A., Drost, M.R., Nicolay, K.: Determination of mouse skeletal muscle architecture using three-dimensional diffusion tensor imaging. Magnetic Resonance in Medicine (2005)

[35] Wang, Z., Petersson, S., Moreno, R., Wang, R.: Anisotropic mechanical properties quantification in skeletal muscle using magnetic resonance elastography and diffusion tensor imaging. Journal of Biomechanics, 112737 (2025)

[36] Chakouch, M.K., Pouletaut, P., Charleux, F., et al.: Quantifying the elastic property of nine thigh muscles using magnetic resonance elastography. PLOS One 10(9), 0138873 (2015)

[37] Felsberg, M., Sommer, G.: The monogenic signal. IEEE transactions on signal processing 49(12), 3136–3144 (2002)

[38] Bridge, C.P.: Introduction to the monogenic signal. arXiv preprint 1703.09199 (2017)

[39] Wachinger, C., Klein, T., Navab, N.: The 2d analytic signal for envelope detection and feature extraction on ultrasound images. Medical image analysis 16(6), 1073– 1084 (2012)

[40] Mei, K., Hu, B., Fei, B., Qin, B.: Phase asymmetry ultrasound despeckling with fractional anisotropic diffusion and total variation. IEEE Transactions on Image Processing 29, 2845–2859 (2019)

[41] Felsberg, M., Sommer, G.: The monogenic signal. IEEE Transactions on Signal Processing 49(12), 3136–3144 (2001)

[42] Alessandrini, M., Basarab, A., Liebgott, H., Bernard, O.: Myocardial motion estimation from medical images using the monogenic signal. IEEE transactions on image processing 22(3), 1084–1095 (2012)

[43] Maltaverne, T., Delachartre, P., Basarab, A.: Motion estimation using the monogenic signal applied to ultrasound elastography. In: 2010 Annual International Conference of the IEEE Engineering in Medicine and Biology, pp. 33–36 (2010). IEEE

[44] Kasai, C., Namekawa, K., Koyano, A., Omoto, R.: Real-time two-dimensional blood flow imaging using an autocorrelation technique. IEEE Transactions on sonics and ultrasonics 32(3), 458–464 (1985)

